# Seroprevalence of Antibodies to SARS-CoV-2 in Healthcare Workers in Non-epidemic Region: A Hospital Report in Iwate Prefecture, Japan

**DOI:** 10.1101/2020.06.15.20132316

**Authors:** Akihiro Nakamura, Ryoichi Sato, Sanae Ando, Natsuko Oana, Eiji Nozaki, Hideaki Endo, Yoshiharu Miyate, Jun Soma, Go Miyata

## Abstract

**Background:** As of June 18, 2020, Iwate is the only one of 47 prefectures in Japan with no confirmed coronavirus disease 2019 (COVID-19) cases. **S**erological survey for COVID-19 antibodies is crucial in area with low prevalence as well as epidemic area when addressing health and social issues caused by COVID-19.

**Methods:** Serum samples from healthcare workers (n = 1,000, mean 40 ± 11 years) of Iwate Prefectural Central Hospital, Iwate, Japan were tested for the prevalence of the severe acute respiratory syndrome coronavirus 2 (SARS-CoV-2) antibodies. Two laboratory-based quantitative tests (Abbott’s and Roche’s immunoassays) and one point-of-care (POC) qualitative test performed simultaneously. Sensitivity and specificity were 100%, 99.6% in Abbott’s immunoassay; 100%, 99.8% in Roche’s immunoassay; 97.8%, 94.6% in Alfa POC test, respectively.

**Results:** The laboratory-based quantitative tests showed positive in 4 of 1,000 samples (0.4%) (95% CI: 0.01 to 0.79): 4/1,000 (0.4%) (95% CI: 0.01 to 0.79) in Abbott; 0/1,000 (0%) in Roche. Positive samples were not detected for both Abbott’s and Roche’s immunoassays. The POC qualitative test showed positive in 33 of 1,000 samples (3.3%) (95% CI: 2.19 to 4.41). There were no samples with simultaneous positive reaction for two quantitative tests and a POC test.

**Conclusions:** Infected COVID-2 cases were not confirmed by a retrospective serological study in healthcare workers of our hospital. The POC qualitative tests with lower specificity have the potential for higher false positive reactions than the laboratory-based quantitative tests in areas with very low prevalence of COVID-19.

## INTRODUCTION

Coronavirus disease 2019 (COVID-19) caused by the severe acute respiratory syndrome coronavirus 2 (SARS-CoV-2)^1,2^ has spread rapidly all over the world and consequently COVID-19 pandemic has seriously affected human health and social life.^3^ In Japan, the first case was confirmed on January 2020^4^ and infected cases have been growing in number across the country. As of May 30, 2020, the cumulative number of them reached 16,650 persons,^4^ but fortunately there have been no officially confirmed cases in Iwate Prefecture.^5^ Because of limited number of real-time reverse transcriptase polymerase chain reaction (RT-PCR) tests to diagnose COVID-19, the accurate prevalence of SARS-CoV-2 infection remains unclear in this area.

Serological survey with SARS-Co-2 antibodies [immunoglobins (Ig) M and/or IgG] would be potentially helpful for understanding of infection status in both individuals and society. Many available point-of-care (POC) antibody tests have been developed and they can be reliable as using in areas with high prevalence of COVID-19.^6^ Although they have the potential for poorly accuracy as using in area with low prevalence, the availability of them in non-epidemic area has not been well studied. Our primary endpoint was to examine the prevalence of COVID-19 of healthcare workers in our hospital in Iwate Prefecture (northeastern area of Japan) where no infected cases have been officially observed. Secondary endpoint was to evaluate the accuracy of the POC antibody tests as compared to the quantitative COVID-19 antibody tests.

## METHODS

### Study design

This was a retrospective study evaluating the prevalence of COVID-19 antibodies in healthcare workers in Iwate Prefecture Central Hospital using currently available immunoassays for detection of antibodies specific to the SARS-CoV-2 virus. Iwate Prefecture with around 1.3 million residents, on the Pacific coast of northeastern Japan, is one of the 47 prefectures in Japan. Iwate Prefecture Central Hospital is located in the city of Morioka, which is located in the center of the prefecture (Fig. 1). Informed consent was obtained from all participants. The study protocol was approved by the ethics committee of the Iwate Prefectural Central Hospital (Approval Number. 343), and carried out according to the principles of the Declaration of Helsinki.

**Fig. 1.**
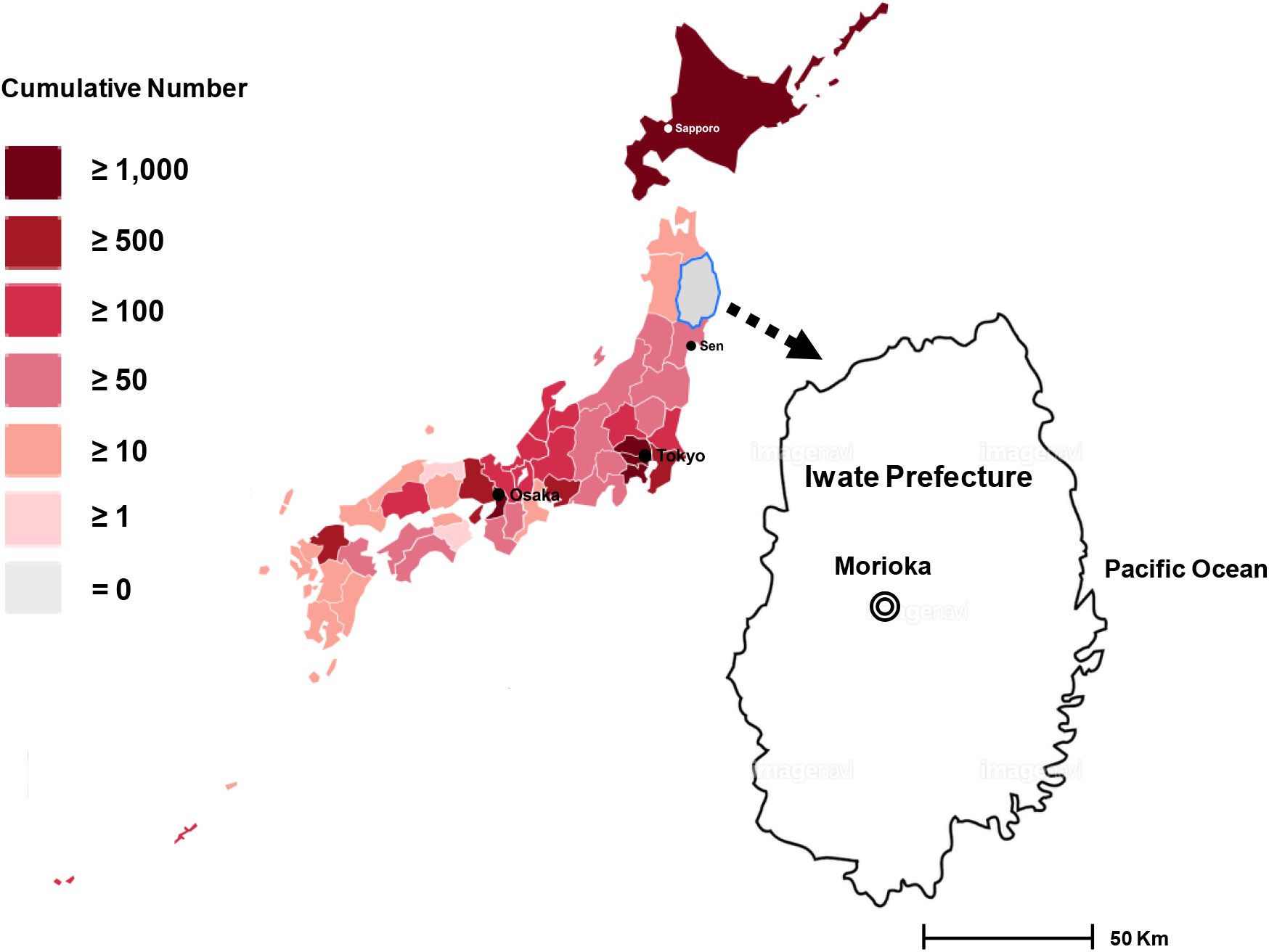
Location of Iwate Prefecture and Iwate Prefectural Central Hospital, and number of COVID-19 cases by prefecture on May 30, 2020 based on the statistics from Ministry of Health and Labor of Japan.^4^

### Study population and antibody tests

A total of 1,302 healthcare workers (physicians, nurses, pharmacists, radiographers, laboratory technicians, and medical office workers) in Iwate Prefectural Central Hospital underwent annual health checkups from May 18 to May 29, 2020, and sera were stored at −20°C until the test in our laboratory. Serum samples (n = 1,000) were analyzed for detection of antibodies to SARS-Co-2 by laboratory-based quantitative and POC qualitative tests. The laboratory-based quantitative tests were performed using Abbott ARCHITECT^®^ SARS-CoV-2 IgG Assay (chemiluminescence microparticle immunoassay; sensitivity: 100%, specificity: 99.6%) (Abbott Laboratories, Abbott Park, IL);^7^ Roche Elecsys^®^ Anti-SARS-CoV-2 RUO Assay (electro chemiluminescence immunoassay; sensitivity: 100%, specificity: 99.8%) (Roche Diagnostics, Basel, Switzerland).^7^ The POC qualitative test was performed using Instant-view^®^ plus COVID-19 Test (lateral flow chromatographic immunoassay; sensitivity: 97.8%, specificity: 94.6%) (Alfa Scientic Designs, Gregg Street Poway, CA). The results were read visually after 10 min. Weak signals for IgM and IgG, together or separate, were considered positive. All tests were conducted at room temperature according to each manufacturer’s instruction.

In this study, a serum sample showing positive for both Abbott’s and Roche’s quantitative immunoassays was considered an infected COVID-19 case. A serum sample showing positive for either Abbott’s or Roche’s immunoassay was a non-infected case. The prevalence of COVID-19 was determined by the number of infected cases divided by the number of measured samples. Continuous variables are expressed as mean ± SD, and categorical variables as numbers and percentages. The 95% CIs for positive rates for the tests were presented in this study.

## RESULTS

Overall, serum samples from 1,000 employees (264 men, 736 women; mean age 40 ± 11 years) were analyzed in this study. Positive reactions were found in 4 of 1,000 (0.4%) (95% CI: 0.01 to 0.79) using Abbott’s quantitative immunoassay; 0 of 1,000 (0%) using Roche’s quantitative immunoassay (Table 1). Presence of COVID-19 cases, which were defined as both positive reaction to SARS-CoV-2 IgG antibody tests using Abbott’s and Roche’s quantitative immunoassays, was not confirmed. The POC qualitative test showed positive in 33 of 1,000 samples (3.3%) (95% CI: 2.19 to 4.41), showing higher rates than those of the laboratory-based quantitative immunoassays. There were no samples with simultaneous positive reactions for two quantitative tests and a POC test (Table 1).

**Table 1.** The results of POC qualitative and laboratory-based quantitative tests for the detection of SARS-CoV-2 antibodies

| Sample number | <u>POC qualitative test</u> |  | <u>Labo-based quantitative test</u> |  |
| --- | --- | --- | --- | --- |
|  | Alfa POC test |  | Abbott Assay | Roche Assay |
|  | IgM | IgG | IgG | IgM + IgG |
| 22 | N | P | N | N |
| 28 | P | N | N | N |
| 32 | P | N | N | N |
| 149 | P | N | N | N |
| 169 | P | N | N | N |
| 227 | P | P | N | N |
| 262 | P | N | N | N |
| 284 | P | N | N | N |
| 286 | P | N | N | N |
| 306 | N | P | N | N |
| 374 | N | P | N | N |
| 382 | P | N | N | N |
| 393 | P | N | N | N |
| 399 | P | N | N | N |
| 409 | P | N | N | N |
| 445 | P | N | N | N |
| 498 | N | P | N | N |
| 545 | P | P | N | N |
| 552 | N | P | N | N |
| 558 | P | N | N | N |
| 571 | N | P | P | N |
| 576 | N | N | P | N |
| 634 | P | N | N | N |
| 694 | N | P | N | N |
| 698 | P | N | N | N |
| 700 | P | N | N | N |
| 726 | N | N | P | N |
| 830 | N | N | P | N |
| 857 | P | N | N | N |
| 882 | N | P | N | N |
| 885 | P | N | N | N |
| 887 | P | N | N | N |
| 904 | P | N | N | N |
| 908 | P | N | N | N |
| 914 | P | N | N | N |
| 926 | P | P | N | N |
| Total number | 33 (IgM or IgG) |  | 4 | 0 |
| Positive rate (95% CI) | 3.3% (2.19-4.41) |  | 0.4% (0.01-0.79) | 0% |

Note. POC, Point-of-Care; SARS-CoV-2, Severe Acute Respiratory Syndrome Coronavirus 2; P, Positive; N, Negative; CI, Confidence Interval.

**Table 2.** Orthogonal comparison of Abbott’s and Roche’s immunoassays

|  | Roche's assay |  |  |
| --- | --- | --- | --- |
|  | Positive | Negative | Total |
| Abbott's assay |  |  |  |
| Positive | 0 (0%) | 4 (0.4%) | 4 (0.0%) |
| Negative | 0 (0%) | 996 (99.6%) | 996 (99.6%) |
| Total | 0 (0%) | 1,000 (100%) | 1000 |

## DISCUSSION

This is the first study to evaluate the positive rate of antibodies to SARS-CoV-2 in officially unidentified epidemic region, Iwate Prefecture in Japan. We qualitatively or quantitatively measured them using three different types of tests, and the results of this study can be summarized as follows. First, serum samples showing positive for both Abbott’s and Roche’s quantitative immunoassays were not detectable. Second, all samples showing positive for the POC qualitative test were considered as false positive reactions without positive responses for Roche’s immunoassay. These results suggest that it seems to be early to judge the accuracy of COCID-2 prevalence using the antibody tests with low specificity even with high sensitivity in the area with very low prevalence

The COVID-19 epidemic of Japan has reached a peak on April 11, 2020 as assessed by daily infected cases, with the number of 714 cases.^4^ After the peak, the number of COVID-19 cases gradually decreased and returned to the early onset levels in January. As of May 30, the total number of confirmed cases was 16,650, and 891 peoples have died,^4^ however infectious cases of Japan are recognized to be relatively small as compared with those of other developed countries. With flattening of epidemic curve as a result of the nation adhering to staying at home and social distancing, the focus has been shifted to widespread testing for COVID-19 antibodies from the RT-PCR test for diagnosis of SARS-CoV-2 infection. Serological tests to detect antibodies against the SARS-CoV-2 are crucial not only for assessment of the immunity of individuals but also for the monitoring the spread of virus and social immunity. In the epicenter showing explosive growth of infected cases such as New York State, the prevalence of COVID-19 has been reported to be as high rate as 10% to 15%,^8^ whereas in the areas where the spread of infection has been minimized such as Santa Clara County, California, US, it has shown to be a lower rate (<1% to 3%).^9^ In Japan, the positive rate of SARS-CoV-2 specific IgG antibodies was 3.3% in 1,000 outpatients visited Kobe City Medical Center General Hospital, Kobe;^10^ 5.9% in 202 blood samples obtained from two community clinics in Tokyo.^11^ According to the Ministry of Health, labor and Welfare, in order to examine the performance of five commercially available test kits, antibody tests were conducted with donated blood collected in Tohoku region in late April, and showed that COVID-19 antibodies were detected in 2 of 500 samples, with a positive rate of 0.4%.^4^ These data and our results are highly suggestive of low prevalence of COVID-19 in Tohoku district compared with that in Tokyo, Osaka, and Kobe city. Our results were also compatible with the results of a nationwide COVID-19 antibodies survey conducted by Ministry of Health and Labor of Japan, showing only 0.03% (1/3,009) positive rate in Miyagi Prefecture next to Iwate.^4^

Iwate Prefecture has been alone among the 47 prefectures in Japan with no reporting COVID-19 cases since Japan has officially recorded its first case on January 2020.^5^ It seems that such situation with no infected cases have never been reported for state-sized regions in other major countries. As of May 30, more than 6,000 inquiries for RT-PCR tests had been made to a local hotline for residents with possible COVID-19 symptoms, however Iwate had conducted only 730 (11.9%) tests which were the lowest level in Japan.^5^ While RT-PCR tests have generally limited to symptomatic patients or patients with known exposure, Iwate had added another hurdle, requiring the tests to be approved by some medical experts until the end of April. Even though it is unclear the reason why Iwate has been skirted SARS-CoV-2, the hospitalized patients with COVID-19 symptoms including coronavirus-infected pneumonia have never been observed so far. COVID-19 cases in neighboring prefectures like Miyagi and Aomori Prefecture^4^ suggest that Iwate may actually harbor SARS-CoV-2 cases but has not detected them due to a low level of tests. Our hospital is located in Morioka, the capital city in the center of Iwate Prefecture, and has given high-quality medical research and service for the people who live in this region. By means of connecting with all other clinics and hospitals by a medical network system, our hospital has played a central role for disease treatment and prevention. On April 7, the state of emergency was declared by the prime minister of Japan,^12^ and the residents in Iwate were required to stay in their home. Because of difficulties in taking a blood sample as part of random sampling from residents in Iwate, we determined the COVID-19 prevalence of healthcare workers in our hospital. Moreover, it would be essential for an assessment of nosocomial infection and provide the underlying data to prevent the spread of infection, potentially leading to a second wave of COVID-19, even though infected cases have never been found in our hospital. We believe that our study will provide the data for informing epidemic models and making public policy decisions.

Recently, several commercial SARS-CoV-2 immunoassays have been developed and available for assessing the immune condition of individuals. They are also essential for control of epidemic infection and making decisions of public policy. In an infectious disease model with very low prevalence, it is recommended to use an antibody test with a very high specificity (perhaps ≥ 99.5%) resulting a high positive predictive value.^13^ In this study, two laboratory-based quantitative tests with a specificity of 99.5% or more (99.6% in Abbott’s assay; 99.8% in Roche’s assay) were used, and positive results were detected in 4 (0.4%) samples in the former while no positive results in the later. As was expected, there were relatively more false positive COVID-19 antibody results in the POC qualitative test, which specificity was relatively low (94.6%). To determine the prevalence of COVID-19 in a given populations, antibody tests with high accuracy and consistent performance are needed not only in Iwate Prefecture but also in other areas in current Japan.

This study has several limitations. First, it was a survey from a single medical institution with a relatively small blood samples. Moreover, the participants in this study were limited to healthcare workers in our hospital, so younger or older people were not enrolled. Second, positive and negative control tests are significant for the full validation of COVID-19 antibodies and the assessment of non-specific binding. In this study, attempts to use the positive or negative control test have presented difficulties because of no identified infected cases and the stay-at-home order. Third, there is still a problem in reliability of antibody tests for COVID-19 and development of accurate tests should be needed. However, they are essential as an epidemiologic tool even in a given population with low prevalence to track progression towards herd immunity for a long time

In conclusion, the positive rate of SARS-CoV-2 antibodies in healthcare workers of our hospital was 0%. Our results suggest the presence of no COVID-19 cases in Iwate Prefecture and the necessity of the antibody tests with high specificity in the areas with very low prevalence.

## Data Availability

Yes

## Acknowledgements

The authors gratefully acknowledge clinical laboratory technologists in Iwate Prefectural Central Hospital for their technical work with blood sampling and measurement.

## Financial support

No financial support was provided relevant to this article.

## Conflicts of interest

All authors report no conflicts of interest relevant to this article.

## Notes

### Competing Interest Statement

The authors have declared no competing interest.

### Funding Statement

This study did not receive any specific grants from funding agencies in the public, commercial, or not-for-profit sectors.

### Author Declarations

The authors declare that they have no competing interests.

